# Determining the indirect costs of suicide in Sweden between 2010 and 2019 - A cost of illness study

**DOI:** 10.1101/2024.09.29.24314575

**Authors:** Daniela Wikström, Camilla Nystrand, Gergö Hadlaczky, Filip Gedin

## Abstract

**Background:** Globally, more than 700 thousand people commit suicide annually. In Sweden, the yearly incidence is between 1000 and 1500 people, which is higher than the global average. Understanding the economic burden of suicide could help highlight the importance and urgency of finding more effective treatments and preventative measures to help people suffering from suicidal thoughts and support studies evaluating the cost-effectiveness of various interventions.

**Method:** This national population-based cross-sectional study estimated the indirect costs associated with all suicides in Sweden between 2010 and 2019. Indirect costs were estimated using the human capital approach. Data regarding average salaries and employment rates were extracted from publicly available data in Sweden. Productivity loss was estimated over two time horizons, the year following the suicide and over a lifetime horizon. Estimations were performed in total numbers for the yearly cohorts as well as per person.

**Results:** Between 2010 and 2019, 1 406 to 1 591 suicides occurred every year in Sweden. In total, approximately 26 500 productive life years are lost every year due to suicide. In 2019, the productivity loss due to all suicides in Sweden was estimated to be 44 million euros over a one-year time horizon, where 10 million euros are direct losses in local and regional authority taxes. Over a lifetime, productivity losses amounted to 935 million euros. The corresponding estimations per person were 37 and 778 thousand euros respectively over a one-year and a lifetime time horizon Over a one-year time horizon, the productivity loss was highest in the older age groups.

**Conclusions:** This study provides valuable insights into the economic burden of suicide on Swedish society. It underlines the potential economic benefits of effective suicide prevention, aligning with previous research highlighting the substantial returns—both monetary and in terms of human well-being—that successful prevention strategies can yield.

## Introduction

More than 700,000 people commit suicide across the world annually (1), and between 1,000 and 1,500 people commit suicide per year in Sweden (2). The incidence of suicide is greater in Sweden than in the rest of the globe, with 10.7 deaths compared to 9 deaths per 100,000 people on average (3). People aged above 65 are more likely to commit suicide, followed by people aged between 45 and 64 years (2). Among adults, men are 2.5 times more likely to commit suicide than women are, and in the 15-to 29-year-old group, suicide is one of the leading causes of death (1). Suicide is classified as a major public health problem, not only because a person dies prematurely but also because of its extensive implications for family members, relatives, friends, and society at large. Although it is well known that mental health issues such as psychotic disorders and mood and anxiety disorders are associated with an increased risk for suicide, a recent meta-analysis estimated that the population attributable risk of mental disorders as a whole was approximately 21% (4). This substantial proportion should not be downplayed, but it is perhaps not surprising that most working-age individuals who die by suicide have no underlying mental health disorder and have an occupation at the time of their death (5).

Previous research has shown that a large part of the costs attributed to suicide do not stem from direct medical costs but rather from costs associated with the loss of labor production (6, 7). However, these cost estimations are not based on complete national registries, introducing uncertainty in the estimations. While Sweden has some of the world’s most comprehensive individual-level registries, it allows for encompassing estimations of the burden of suicide.

Understanding the economic burden of suicide could help highlight the importance and urgency of finding more effective treatments and preventative measures to help people suffering from suicidal thoughts and support studies evaluating the cost-effectiveness of various interventions. In addition, how the economic burden is spread across different actors in society is important to policy makers and for budget allocation because it provides arguments for the potential economic benefit for different actors to invest in preventive measures. The aim of this study is to estimate the economic burden related to indirect costs that suicide has imposed on Swedish society between 2010 and 2019.

## Methods

This national population-based cross-sectional study estimated the indirect costs associated with all suicides in Sweden between 2010 and 2019. Indirect costs were estimated using the human capital approach. This study was conducted in accordance with the STROBE reporting guidelines (see Supplementary Table 1). All data used in the study have been aggregated, publicly available data. Hence no ethical approval or consent has been necessary.

### Data collection

Data regarding the number of suicides committed in Sweden per year were acquired from the Swedish National Cause of Death Registry, which is held by the Swedish National Board of Health and Welfare. The data were accessed for research purposes from 2010 to 2019. Authors did not have access to information that could identify individual participants during or after data collection. To approximate the true incidence of suicides in Sweden, suicide deaths classified as ICDs X60-X84/E950-E959 were combined with so-called uncertain suicides, i.e., ICD Y10-Y34/E980-E989, between 2010 and 2019. The data were divided into yearly cohorts based on the year when the suicide occurred. Each yearly cohort was further subgrouped based on sex and age.

The parameters included in the calculations for productivity loss are outlined in Table 1. These included the average salary in Sweden retrieved from Statistics Sweden and data on average employment, social fees, the pension rate and vacation compensation. Furthermore, Table 1 displays the division of income taxes paid to the regional and local authorities.

**Table 1.**
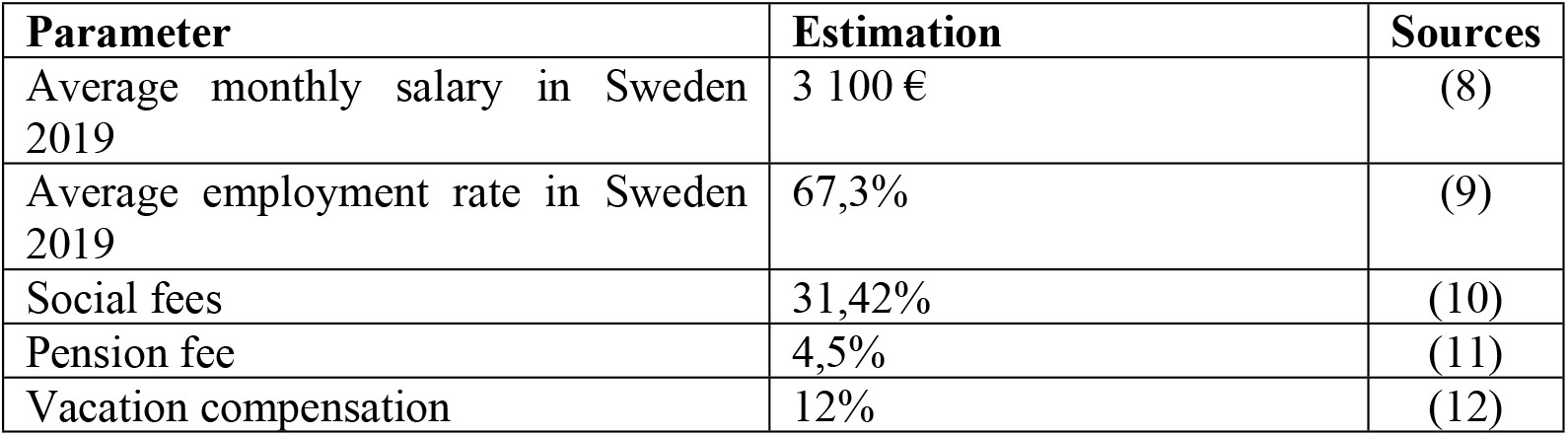
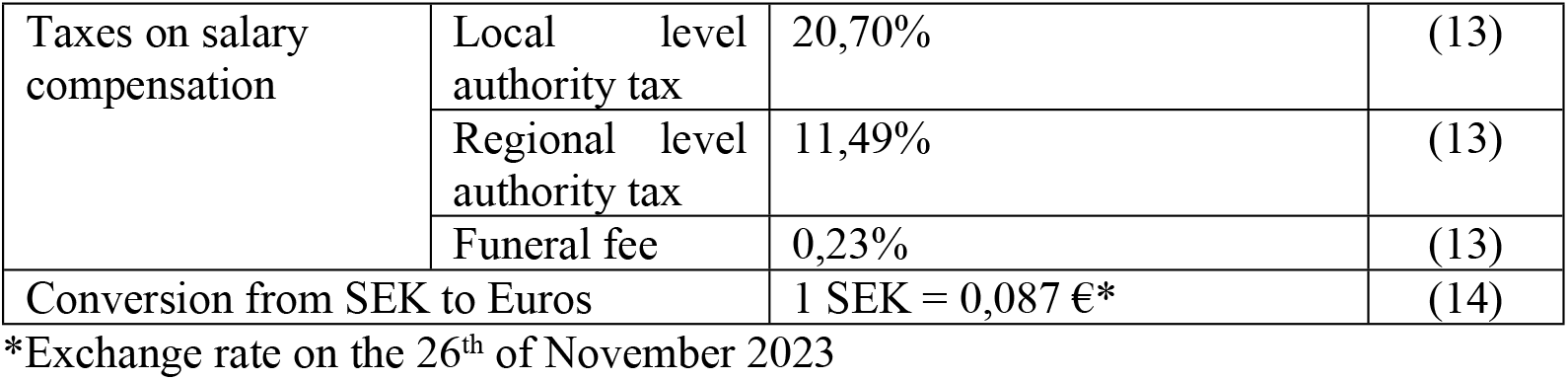
Parameters included in the calculations.

| Parameter | Estimation | Sources |
| --- | --- | --- |
| Average monthly salary in Sweden 2019 | 3 100 € | (8) |
| Average employment rate in Sweden 2019 | 67,3% | (9) |
| Social fees | 31,42% | (10) |
| Pension fee | 4,5% | (11) |
| Vacation compensation | 12% | (12) |

|  |  |  |  |
| --- | --- | --- | --- |
| Taxes on salary compensation | Local level authority tax | 20,70% | (13) |
|  | Regional level authority tax | 11,49% | (13) |
|  | Funeral fee | 0,23% | (13) |
| Conversion from SEK to Euros |  | 1 SEK = 0,087 €* | (14) |

\*Exchange rate on the 26<sup>th</sup> of November 2023

### Cost analysis

#### Productivity loss

The human capital approach was used to estimate the productivity loss that occurs due to premature death following completed suicide. Years of productive life lost were estimated by subtracting the age when the suicide occurred from the retirement age in Sweden and including the average mortality rate for each age (probability of surviving another year).

Productivity loss was calculated over two different time horizons, the year following the death and over the lifetime. Estimations were performed in total numbers for the yearly cohorts as well as per person. Productivity loss was further estimated over five different age groups, 18-24, 25-34, 35-44, 45-54 and 55-67, both over a one-year time horizon and over a lifetime. The parameters included in the estimations were the average employment rate in Sweden, the average monthly salary, social fees (31.42% of the monthly salary), vacation compensation (12% of the monthly salary) and the pension fee paid by employers (4.5% of the monthly salary); see Table 1. All calculations were performed on a yearly basis.

Due to premature death and loss of productivity, individuals are unable to contribute to the tax system at large. Taxes in Sweden are divided into state-, regional- and local-level authorities. Most of these taxes are paid by individuals through salary compensation and value added tax (VAT) on goods and services purchased and by employers through social and pension fees. Tax loss (part of the total indirect cost) was estimated as the percentage of the monthly income that would have gone to taxes, with a regional tax of 11.49% of the monthly income and a local authority tax of 20.70% of the monthly salary.

#### Discounting, Economic Growth, and Inflation

For the lifetime horizon, future costs were discounted at an annual rate of 3%, which is recommended for health economic evaluations in Sweden (15). This discount rate accounts for the time preference of money, reflecting the principle that costs incurred in the future are less valuable than costs incurred today. Additionally, future economic growth rates and inflation were considered by adjusting the average salary growth rate based on historical data from Statistics Sweden. The average annual growth rate of salaries was assumed to be 2%, and the inflation rate was assumed to be 1.5%, based on historical trends.

#### Net vs. Gross Salary

The salary data used in this study represent gross salaries, which include all taxes and social fees. The salaries were adjusted for age-specific variations. This approach provides a comprehensive estimate of the economic impact of suicide by capturing the total earnings lost, including the portions that would have been paid as taxes and social contributions.

#### Sensitivity analysis

In the sensitivity analysis, the employment rate was varied to 40 percent, 50 percent, 60 percent, 70 percent, and 80 percent to account for the possible difference in productivity for this specific cohort.

## Results

Between 2010 and 2019, 1 406 to 1 591 suicides occurred every year in Sweden. The average age of people who commit suicide during these years was approximately 50 years. In total, approximately 26 500 productive life years were lost every year due to suicide (supplementary table 2).

### Total cost

Between 2010 and 2019, the total cost of suicide related to indirect costs varied between 32 million and 44 million euros over a one-year horizon and between 795 million and 935 million euros over a lifetime horizon. The indirect costs per person varied between 28 thousand and 37 thousand euros over a one-year horizon and between 708 thousand and 778 thousand euros over a lifetime horizon (Table 2).

**Table 2.** Total forgone costs due to suicide over a one-year time horizon and a lifetime horizon.

| Year | Productivity losses |  | Productivity losses per person |  |
| --- | --- | --- | --- | --- |
|  | 1 year | Lifetime | 1 year | Lifetime |
| 2010 | 32 000 000 € | 795 000 000 € | 28 000 € | 708 000 € |
| 2011 | 33 000 000 € | 830 000 000 € | 30 000 € | 758 000 € |
| 2012 | 37 000 000 € | 865 000 000 € | 31 000 € | 725 000 € |
| 2013 | 38 000 000 € | 926 000 000 € | 31 000 € | 760 000 € |
| 2014 | 38 000 000 € | 909 000 000 € | 32 000 € | 769 000 € |
| 2015 | 38 000 000 € | 891 000 000 € | 32 000 € | 743 000 € |
| 2016 | 38 000 000 € | 834 000 000 € | 34 000 € | 751 000 € |
| 2017 | 41 000 000 € | 883 000 000 € | 35 000 € | 769 000 € |
| 2018 | 44 000 000 € | 926 000 000 € | 37 000 € | 769 000 € |
| 2019 | 44 000 000 € | 935 000 000 € | 37 000 € | 778 000 € |

*Exchange rate of Swedish krona to Euros on the 26<sup>th</sup> of November 2023. 1 SEK = 0,087 €*

### Age groups

The productivity loss between different age groups was estimated (supplementary table 3) both in total and per person. Over a one-year period, the highest total and per-person costs were found for the 45-to 54-year-old age group, followed by the 35-to 44-year-old age group. The cost per person for the 45-to 54-year-old age group ranged between 54 thousand euros and 56 thousand euros. The lowest costs are found in the 18-24 age group; in this age group, the costs vary between 17 thousand euros and 19 thousand euros per person.

Over a lifetime horizon (Table 3), the highest cost is found for the 18-24 age group per person. For this group, the costs vary by approximately 20 million SEK. The lowest costs are found in the 55-to 67-year-old age group, where the costs vary by approximately 3 million SEK.

**Table 3.** Total cost per person for different age groups over a lifetime horizon.

| Age group | Productivity loss over a lifetime | Productivity loss per person over a lifetime |
| --- | --- | --- |
| 18-24 | 227 000 000 € | 1 600 000 € |
| 25-34 | 411 000 000 € | 1 500 000 € |
| 35-44 | 262 000 000 € | 1 200 000 € |
| 45-54 | 192 000 000 € | 720 000 € |
| 55-67 | 72 000 000 € | 280 000 € |

### Loss of taxes 2019

As part of the indirect costs of suicide in 2019, the losses in taxes for different sectors amounted to approximately 6.4 million euros for the local authorities and 3.6 million euros for the regional authorities. The lost social fees that would have been transferred to the state for the cohort that passed away in 2019 amount to 14.5 million euros.

### Sensitivity analysis

In the sensitivity analysis, the employment rate varied between 40 and 80%. The average employment rate in Sweden is approximately 67 percent. In 2019, the indirect costs over a one-year period varied between 20 thousand euros and 40 thousand euros per person. Over a lifetime, the costs varied between 428 thousand and 856 thousand euros. In the different age groups, the indirect cost (per person) over a one-year period varied between 15 thousand – 31 thousand euros (age group 18-24), 18 thousand – 37 thousand euros (age group 25-34), 22 thousand – 44 thousand euros (age group 35-44), 23 thousand – 46 thousand euros (age group 45-54) and 22 thousand – 45 thousand euros (age group 55-67). Over a lifetime horizon, the indirect costs due to suicide (per person) varied between 734 thousand – 1.5 million euros (age group 18-24), 655 thousand – 1.3 million euros (age group 25-34), 507 thousand – 1 million euros (age group 35-44), 323 thousand – 647 thousand euros (age group 45-54) and 131 thousand – 262 thousand euros (age group 55-67).

## Discussion

This study provides valuable insights into the economic burden of suicide on Swedish society. The findings reveal substantial financial implications, with suicide accounting for significant amounts in terms of both one-year and lifetime estimates. In 2019, the total productivity losses amounted to 44 000 000 € yearly. Some of these losses fall on the state (14.5 million euros), regional (3.6 million euros) and local (6.4 million euros) authorities due to tax losses.

The costs per person, though varying across different studies, offer a comparative perspective. A study from Finland estimated substantially lower costs per person over a lifetime perspective than reported in this study (322,928 euros compared with 729,575 euros). The reasons for this are the different methods used in the estimation. We estimated indirect costs based on loss of production, whereas Solin and colleagues measured costs only in terms of lost tax returns in Finland [7]. In contrast, an Australian study which estimated indirect costs of suicide for individuals ages 24 and younger, found a similar yearly indirect cost, 1 700 000 euros, compared to our estimates of 1 600 000 euros for individuals aged 24 and younger [15].

This study underlines the potential economic benefits of effective suicide prevention, aligning with previous research highlighting the substantial returns, both monetary and in terms of human well-being, that successful prevention strategies can yield. The economic foundation provided by this study can provide powerful support for knowledge-based decision-making as well as for the development and evaluation of preventive measures.

The focus on costs associated with loss of productivity excludes other substantial costs related to suicide, such as emergency response, medical care, family members’ incapacity for work and/or potential treatment costs related to complicated grief and other disorders of relatives, costs related to the transport system (e.g., in railway, subway and motorway suicides) and property damage. Both Kinchin et al. and Solin et al. included costs related to direct medical or emergency expenditures (7, 16). However, the direct cost only accounted for a fraction of the total cost of suicide, and the other unrelated costs are also likely to be only a fraction of the total costs related to suicide. In the Kinchin et al. study, health-care costs during suicide-related emergencies only accounted for 0.01% of the total cost, and in the study by Solin et al., the productivity loss from deceased people accounted for approximately 70%. Similarly, a study by Segar et al. estimating the costs of suicides in France found that the direct costs of completed suicides are very small compared to indirect costs, with direct costs accounting for only 0.4%, highlighting the significant economic burden of productivity losses and other societal impacts (17).

In this study, we have chosen to focus exclusively on completed suicides for several reasons. Firstly, completed suicides are clearly defined and recorded in national databases, providing us with accurate and reliable data, which minimizes the risk of inaccuracies and uncertainties in our analyses. Secondly, completed suicides have a direct and measurable economic impact on society, including lost productivity and tax revenues. By focusing on these, we can clearly demonstrate the economic burden and the need for effective preventive measures. Thirdly, policymakers and decision-makers require concrete and specific data to make informed decisions about resource allocation and the development of preventive strategies. By focusing on completed suicides, we provide information needed to prioritize preventive interventions that may save lives. By limiting our scope to completed suicides, we can conduct a more focused and in-depth analysis. Lastly, previous studies in this field have also focused on completed suicides (7, 16). By doing the same, the results are comparable to previous research and contribute to a more coherent and comparable knowledge base.

This study has several strengths. First, it uses data from high-quality nationwide registries, which eliminates selection bias and ensures that all cases of suicide in Sweden are included in the dataset. Second, the complete dataset allows for a comprehensive analysis of the data, which is a major strength of the study. Third, the assumptions made in the study are conservative, which means that the results are probably an underestimation of the total cost burden.

The study also has some limitations. One limitation is the decision to only include individuals aged 18-67 years, based on presumed employment, which raises concerns about the exclusion of a significant portion of the population. The age for retirement in Sweden is 67, and it is common to assume that income and tax-generating productivity are subsequently reduced to zero. However, this group, particularly the 85+ group, has the highest suicide risk among all age groups, and most individuals who retire in Sweden continue to contribute to unpaid work. However, the consequence of using the human capital approach for estimating the costs of suicide is the exclusion of this group from the analysis. Another limitation is the precision of our estimates. The Swedish nation-wide registers used as data inputs were on an aggregate level; hence, the employment and salary for each individual were not available, and average estimates had to be used. These limitations should be kept in mind when interpreting the findings of this study. Finally, we included both certain and uncertain suicides in the estimation of productivity losses. Some of the uncertain suicides may not have been suicides, which may have led to an overestimation of the total indirect costs of suicide.

## Conclusion

This study sheds light on the economic burden of suicide in Sweden, emphasizing the need for comprehensive preventive strategies. to the study provides a nuanced understanding of the economic implications of completed suicides, laying the groundwork for future research and policy discussions in the realm of suicide prevention.

## Data Availability

The data are available from the authors upon reasonable request and with the permission of National Centre for Suicide Research and Prevention (NASP).

## Ethics approval and consent to participate

Not applicable

## Consent for publication

Not applicable

## Authors’ contributions

DW, CN, FG, and GH Performed the analyses, collected the data, conceived and designed the analysis and wrote the paper.

## Acknowledgements

This research was partly funded by grants from the Swedish research council for health working life and welfare (2022-00922).

## References

1. Organization TWH. Suicide 2024 [updated 20230828; cited 2024 30 January]. Available from: https://www.who.int/news-room/fact-sheets/detail/suicide.

2. lll-Health NCfSRaPoM. Interpreting suicide data 2023 [updated 06212023; cited 2024 January 30]. Available from: https://ki.se/en/nasp/interpreting-suicide-data.

3. Dattani S, Rodés-Guirao L, Ritchie H, Roser M, Ortiz-Ospina E. Suicides 2023. Available from: https://ourworldindata.org/suicide.

4. Too LS, Spittal MJ, Bugeja L, Reifels L, Butterworth P, Pirkis J. The association between mental disorders and suicide: A systematic review and meta-analysis of record linkage studies. J Affect Disord. 2019;259:302–13.

5. Matilla Santander N, Blazevska B, Carli V, Hadlaczky G, Linnersjö A, Bodin T, et al. Relation between occupation, gender dominance in the occupation and workplace and suicide in Sweden: a longitudinal study. BMJ Open. 2022;12(6):e060096.

6. Shepard DS, Gurewich D, Lwin AK, Reed GA, Jr., Silverman MM. Suicide and Suicidal Attempts in the United States: Costs and Policy Implications. Suicide Life Threat Behav. 2016;46(3):352–62.

7. Solin P, Tamminen N, Seppänen A, Partonen T. Calculation of Costs Related to Death by Suicide in Finland. Social Sciences. 2022;11(10):468.

8. Swedish National Mediation Office. Löneskillnaden mellan kvinnor och män 2019 : vad säger den officiella lönestatistiken? Stockholm: Medlingsinstitutet; 2020.

9. SCB. Arbetskraftsundersökningarna (AKU) 2019 [Elektronisk resurs]. Stockholm: Statistiska centralbyrån; 2020.

10. Skatteverket. Arbetsgivaravgifter 2019 [Available from: https://www.skatteverket.se/foretag/arbetsgivare/arbetsgivaravgifterochskatteavdrag/arbetsgivaravgifter.4.233f91f71260075abe8800020817.html.

11. Swedish Tax Agency. Anställd inom kommun och region: Swedish Tax Agency,; 2023 [Available from: https://www.pensionsmyndigheten.se/forsta-din-pension/tjanstepension/anstalld-inom-kommun-och-region.

12. Labour Mo. The Swedish annual leave act. Stockholm: Ministry of Labour; 1985.

13. Statistics Sweden. The total municipal tax rate increases next year: Statistics Sweden, ; 2023 [Available from: https://www.scb.se/en/finding-statistics/statistics-by-subject-area/public-finances/local-government-finances/local-taxes/pong/statistical-news/municipal-taxes-2024/.

14. European central bank (ECB). Euro foreign exchange reference rates: ECB; 2024 [Available from: https://www.ecb.europa.eu/stats/policy_and_exchange_rates/euro_reference_exchange_rates/html/eurofxref-graph-sek.en.html.

15. (TLV) T-ol. Allmänna råd om ekonomiska utvärderingar (TLVAR 2017:1). 2017.

16. Kinchin I, Doran CM. The Cost of Youth Suicide in Australia. Int J Environ Res Public Health. 2018;15(4).

17. Segar LB, Laidi C, Godin O, Courtet P, Vaiva G, Leboyer M, et al. The cost of illness and burden of suicide and suicide attempts in France. BMC Psychiatry. 2024;24(1):215.

